# Florida Doulas’ Perspectives on their Role in Reducing Maternal Morbidity and Health Disparities

**DOI:** 10.1101/2023.04.19.23288758

**Authors:** Janelle Applequist, Roneé Wilson, Megan Perkins, Charlette Williams, Ria Joglekar, Richard Powis, Angela Daniel, Adetola F. Louis-Jacques

## Abstract

**Introduction:** Maternal mortality rates continue to rise in the United States. Considerable racial disparities exist, as Black women are 2-3 times more likely to die from pregnancy-related complications than White women. Doulas have been associated with improved maternal outcomes. This study aimed to 1) investigate Florida doulas’ perspectives on severe maternal morbidity/mortality, related inequities, and their influence on these areas as well as 2) identify opportunities for actionable change.

**Methods:** This qualitative study included seven online, in-depth interviews and seven focus groups with doulas (*n* = 31) in the state of Florida. Interview and focus group guides aimed to investigate how doulas perceive their role in the context of a) maternal morbidity and b) health disparities/inequities.

**Results:** Doulas associated maternal morbidity and health disparities with Black pregnant people, attributing racism as a major contributor. Doulas identified their role in mitigating this problem as one that involves: providing positive social surveillance and emotional support, equipping clients with access to education and resources, and championing for advocacy in healthcare settings. Actionable steps utilizing the social ecological model and reproductive justice framework are provided.

**Discussion and Health Equity Implications:** Doulas perceived their role as integral to mitigating maternal morbidity and maternal health disparities, particularly in the context of race. Equitable access to doulas for low-income and or minoritized populations may be one key strategy to improve maternal health equity.

## Introduction

Pregnant people in the US have not reaped the benefits of the decline in worldwide maternal mortality (MM) rates.^1^ The national MM rate rose to 32.9 deaths per 100,000 live births in 2021, compared to a rate of 23.8 in 2020.^2^ Even after controlling for education and socioeconomic status, Black birthing people remain at the highest risk for MM and severe maternal morbidity (SMM), with a MM rate 2.6 times the rate of non-Hispanic White birthing people.^2-5^ Comparable trends persist in Florida.^6, 7^ Many multilevel factors may explain worsening US maternal outcomes. A systematic review^8^ on social determinants of health reported that insurance status (i.e., covered by Medicaid or not at all), income level, and lower education are associated with higher risk of maternal morbidity and mortality, in additional to demographic characteristics such as Black race and Hispanic ethnicity.

Doula support has been associated with improved birth outcomes including fewer cesareans, shorter labors, less birth complications, higher breastfeeding rates, higher rates of birth experience satisfaction.^9-12^ Furthermore, the inclusion of community-based, culturally competent doula support has been associated with fewer maternal complications among marginalized women facing higher rates of MM/SMM.^11-15^ Historically, doulas were utilized in White, middle/higher class spheres,^16^ and low income, as well as other marginalized, populations had limited access to these services.

Despite popular discourse on how doula services may mitigate MM/SMM and related inequities,^17-20^ there are limited data assessing the perspectives of doulas on their perceived role in these areas. Our study aims to 1) investigate Florida doulas’ perspectives of MM/SMM and health inequities and their influence on these areas and 2) identify opportunities for actionable change in the context of maternal morbidity and maternal health disparities.

## Methods

Online, in-depth interviews and focus groups were conducted with 31 doulas serving Florida families to gather rich data related to their lived experiences in the profession. Interviews and focus groups were chosen for purposes of data triangulation, as focus groups can lead to conformity of one opinion. Therefore, interviews served as a form of data checking to ensure that thematic representations were sound and not being led by focus group dynamics. Seven interviews and seven focus groups were conducted.

Interview and focus group guides were developed by all team members and were informed by the social ecological model (SEM; Figure 1). The SEM is based on the theory that an individual’s behavior is integrated in a dynamic network of intrapersonal characteristics, interpersonal processes, institutional factors, community features and public policy.^21^ Secondarily, these guides were developed with the acknowledgement that multiple factors influence health inequities, which is one of the tenets of reproductive justice. Reproductive justice advocates for listening to birthing persons, recognizing autonomy, amplifying their voices to oppose oppression and to regain reproductive control.^22, 23^ For this study, we focused on how doulas interpret their role within the context of the SEM.

**Figure 1.**
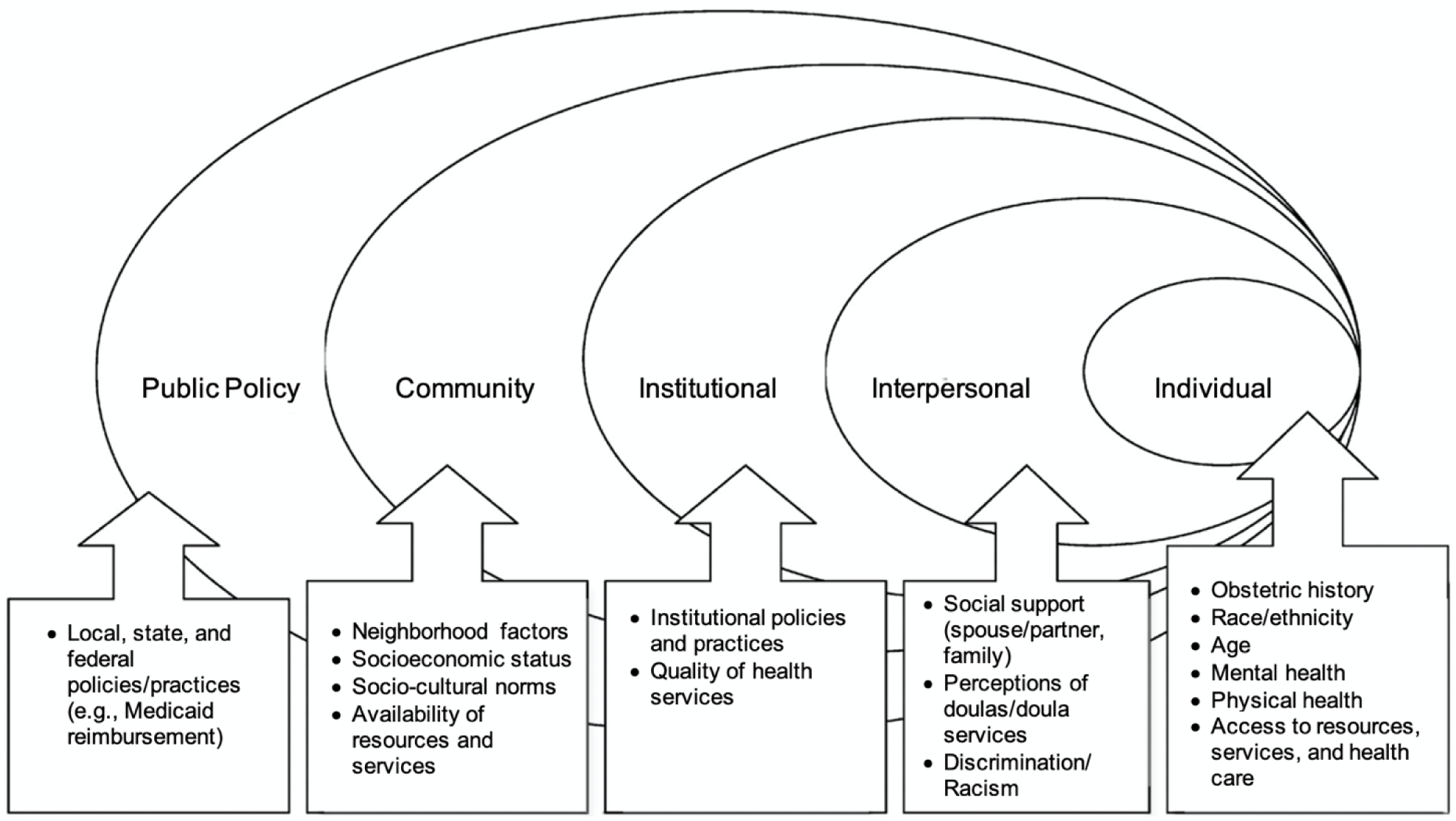
Conceptual Model - Social Ecological Model (SEM) A conceptual model of our study, based on the social ecological model, which considers how a variety of factors influence a birthing person’s pregnancy journey.

The study was approved as exempt by the University of South Florida and the University of Florida Institutional Review Boards. Convenience and snowball sampling practices were used to recruit doula participants. Flyers advertising the study were shared with community partners and distributed within their networks. The time interval for recruitment and data collection was approximately four months (April-July 2022). Interested participants completed an online pre-screening questionnaire which was used to determine eligibility (e.g., labor and delivery doula experience in Florida, served at least two families in the past year). After providing consent, doula participants participated in an interview or focus group lasting approximately 60 minutes. Participants were compensated for their time with a $40 digital gift card of their choice to Target, Amazon, or Wal-Mart. Interviews and focus groups were recorded via Zoom and auto-transcribed via artificial intelligence technology.

During interviews and focus groups, doulas were asked to provide the first word that came to mind upon hearing the phrase ’maternal morbidity,’ to discuss health disparities, and were asked how they felt their skill sets may help to reduce such disparities. The subject of race or racism was not raised by the interviewer. To identify actionable steps for solutions, doulas were asked how they would address the problems they were articulating in their sessions.

Team members cleaned transcripts to ensure accuracy. All team members reviewed transcripts for an inductive approach toward codebook generation, following Campbell et al.’s method for coding interview data.^24^ All team members performed open coding of all transcripts, where a close line-by-line reading of all data occurred, allowing for meaningful words, units, and contexts to emerge.^25^ Axial coding was performed next, all team members met to combine initial open codes into broader themes, where relevant attributes could be further explored.^26^ Themes generated reached theoretical saturation (each thematic category became operationalized with no novel information presented in subsequent interviews or focus groups), which is a significant component of qualitative research.^27^ The first author unitized 10% of the sample of transcripts, and following Campbell et al.’s approach for achieving intercoder agreement, the first and third authors independently coded these data.^24^ According to Miles and Huberman’s accepted levels for reliability,^28^ the overall intercoder agreement was “very good” at a level of 92.2%. All data were then coded by the first author in NVivo, a qualitative data analysis software, to assess thematic patterns.

## Results

### Demographics

Most doulas interviewed identified as female. Most participants identified as birth, post-partum, and full spectrum doulas. Most (69%) of the doulas who participated were between 35 and 54 years of age. Three-quarters (75.9%) self-reported their race as White, 17.2% self-reported their race as Black, and 6.9% self-reported their race as other. One-quarter of participants (27.6%) self-reported as Hispanic/Latina. While more than half (58.6%) of the doula participants reported an income greater than $50,000 per year, nearly all (93.1%) reported serving families who may qualify for government assistance due to economically disadvantaged circumstances. These families may be eligible for Medicaid, WIC, or free/reduced lunch.

To provide depth and breadth while privileging doula responses, summaries of thematic findings, relevant SEM level, and relevant participant quotes are provided in Table 1. The names of the doulas that are quoted in Table 1 have been changed to pseudonyms to protect their privacy. In response to the first prompt, **theme one** found that doulas overwhelmingly (98%) associated *maternal morbidity* with *Black* or *African American pregnant people* without the interviewer prompting for racial disparities. A word cloud was created to visually represent the saturation of responses, with phrases provided most often appearing in larger font (Figure 2).

**Figure 2.**
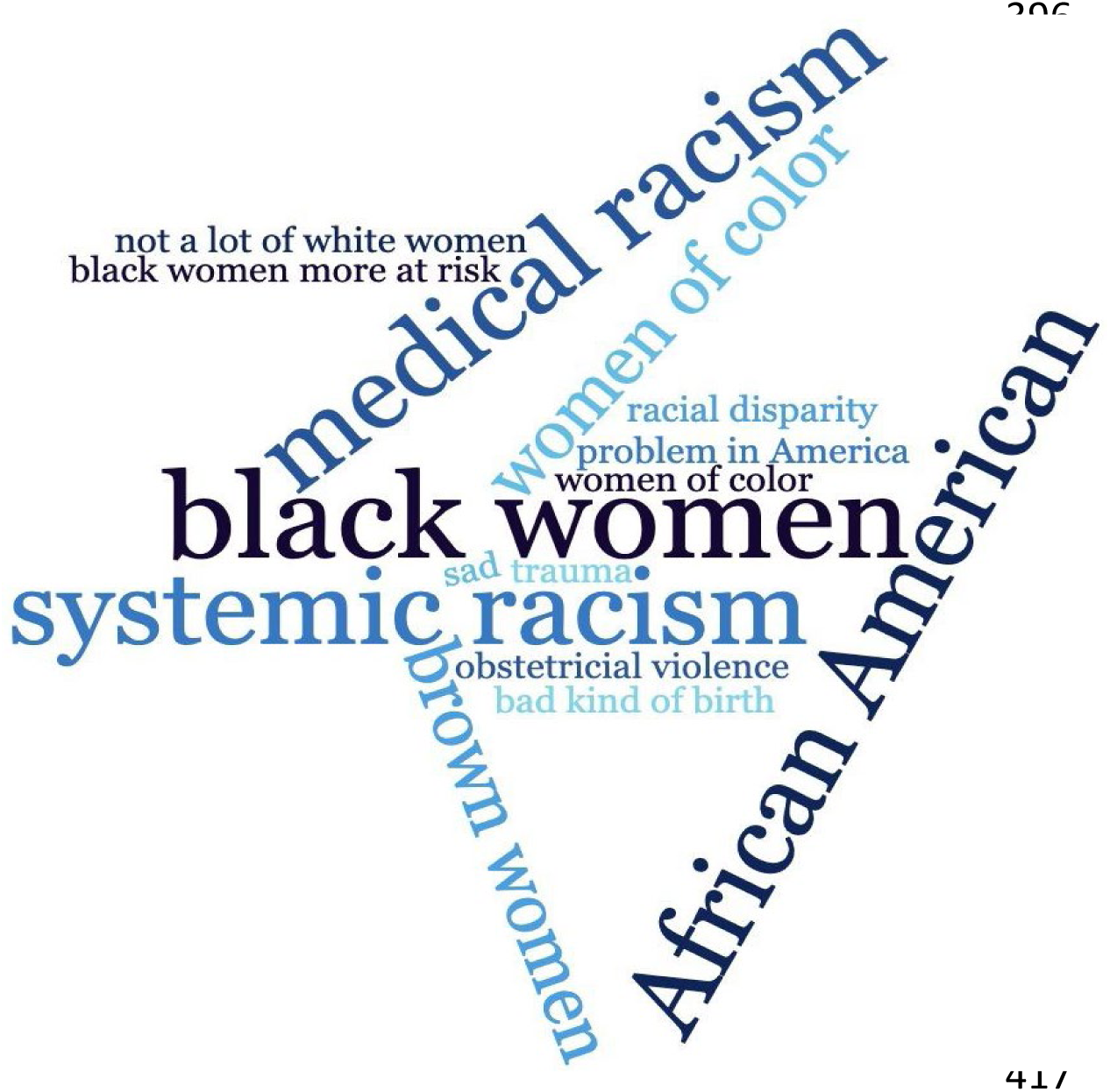
Word Cloud Representation. Doula Responses to “When you hear the phrase ‘maternal morbidity,’ what is the first word that comes to mind?”

**Table 1.**
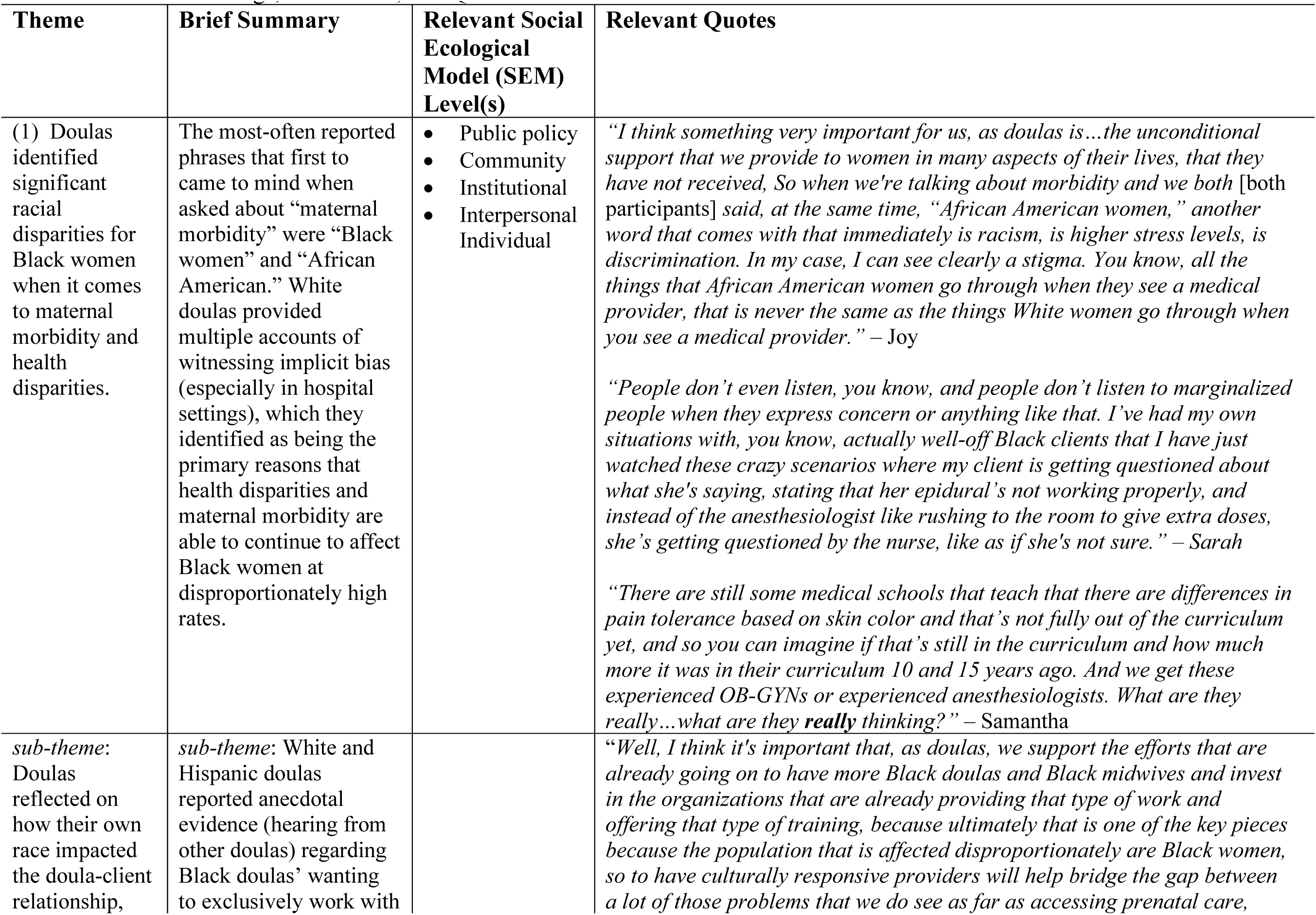
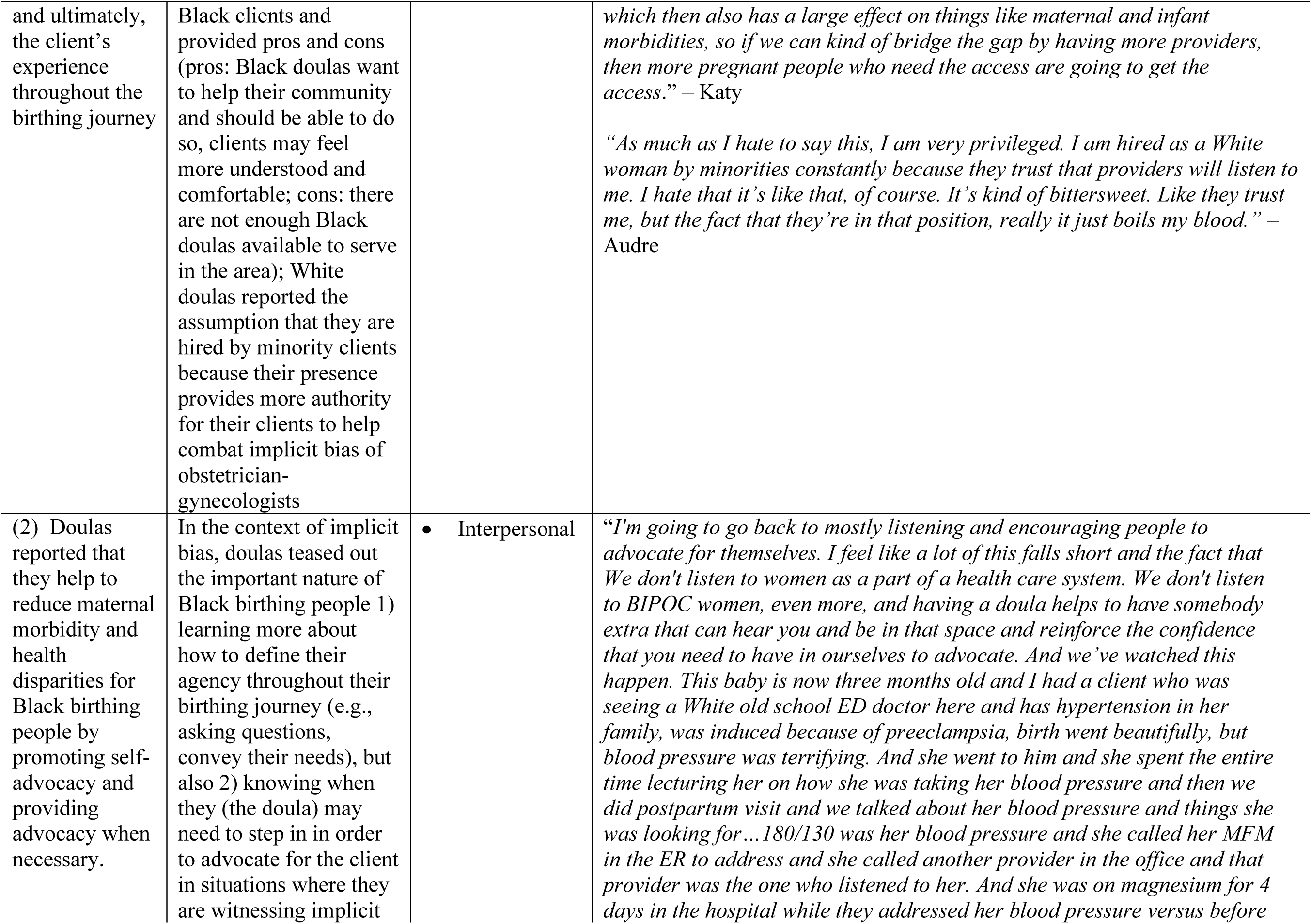
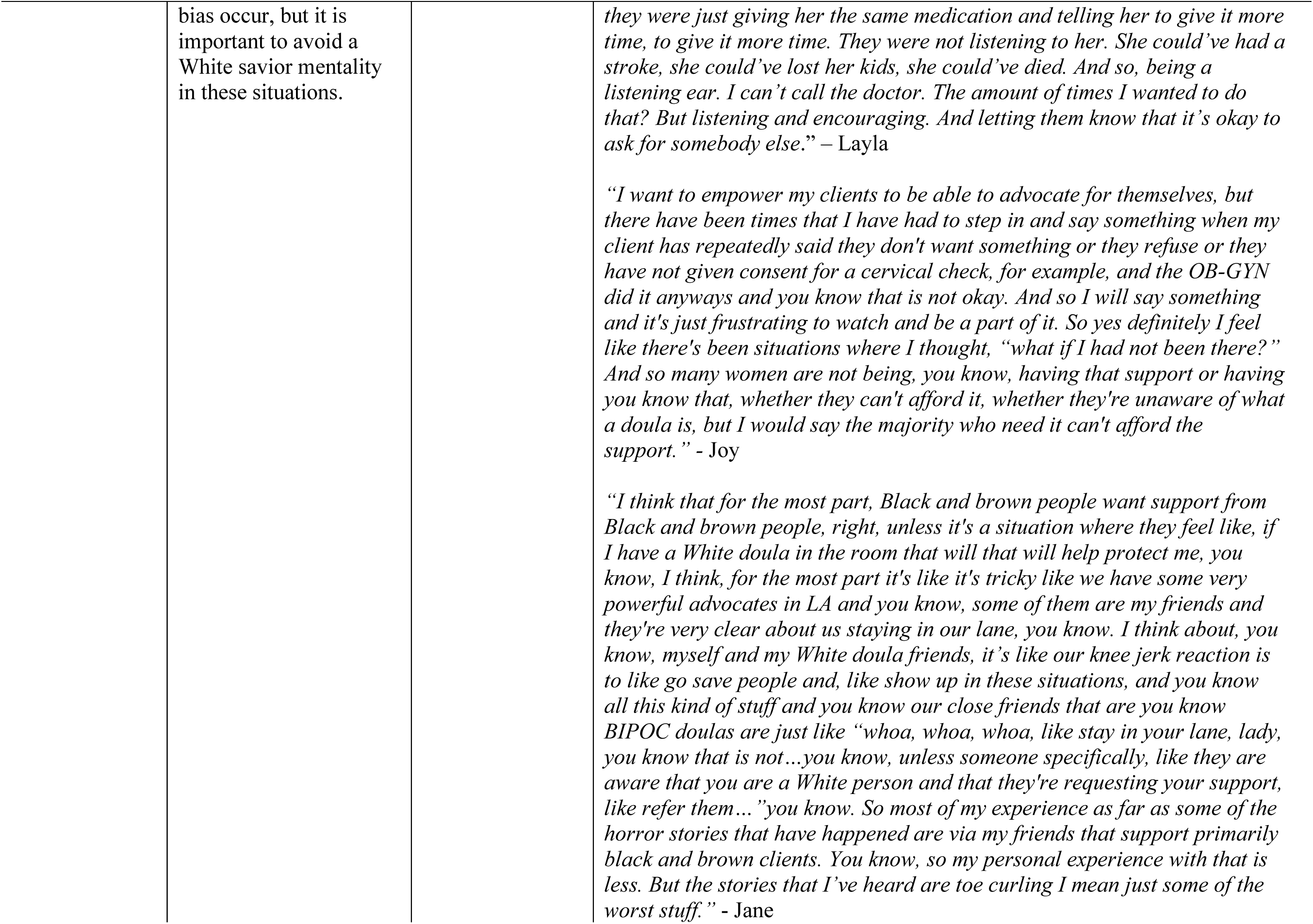
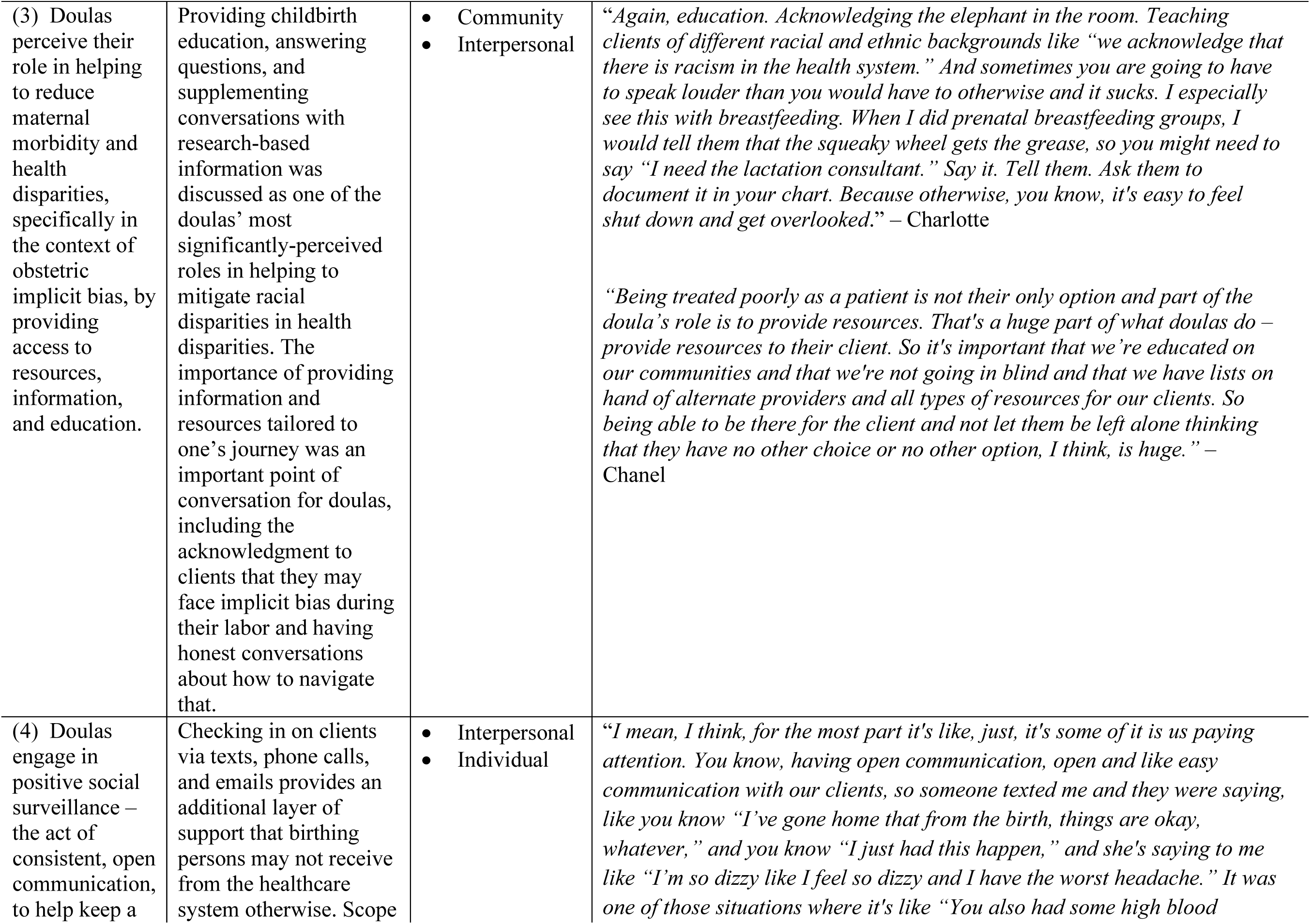
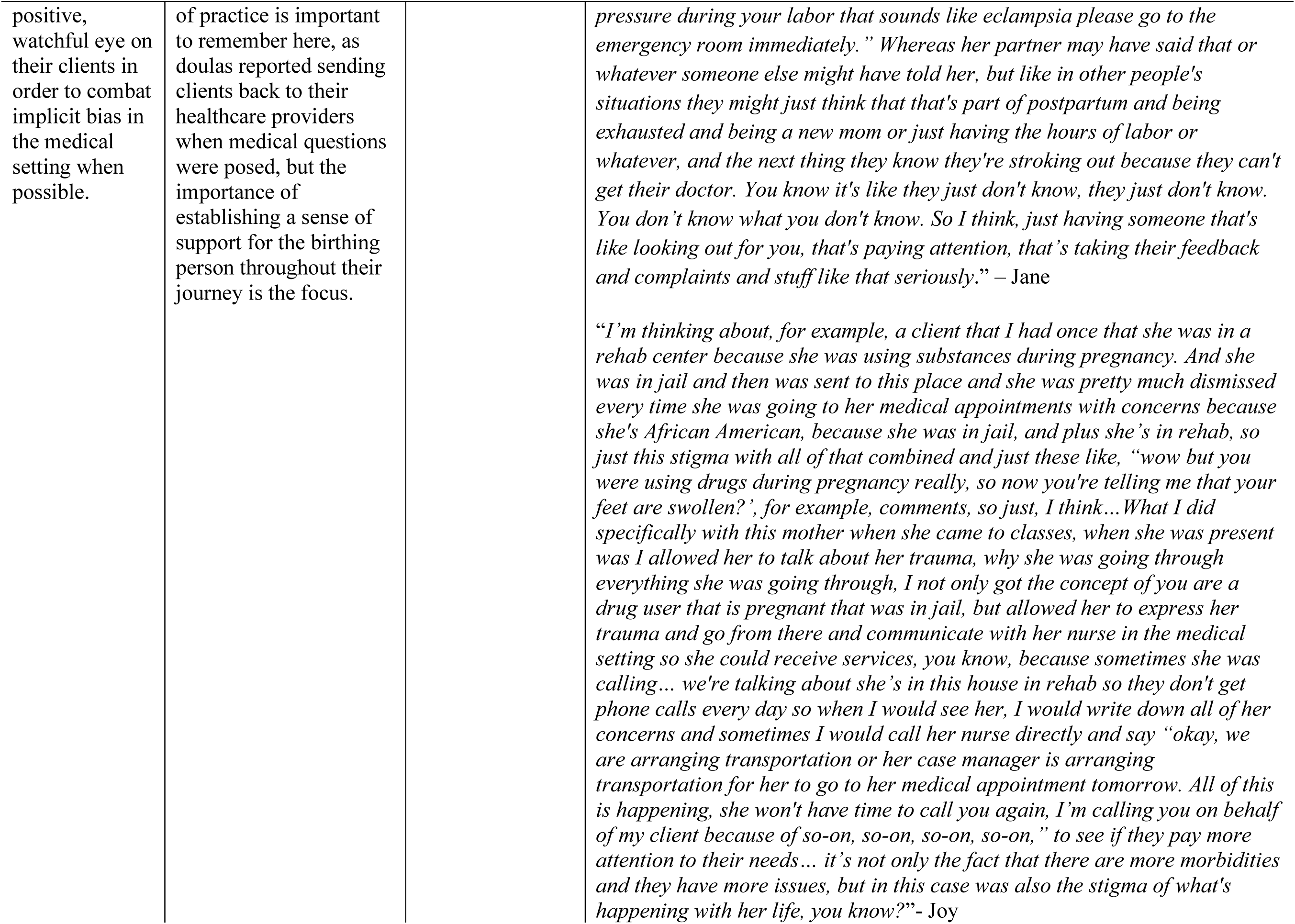
Thematic Findings, Summaries, and Quotes.

Frustration and sadness were expressed in the conversations surrounding this subject, and as dialogue continued, important conversations began to occur surrounding the race of doulas and their clients. A few doulas reported hearing that Black doulas preferred working with Black clients. Positive sentiment regarding this idea featured the thought that members of a community could connect with their clients, understand them, and educate them in more powerful ways. Other ideas posed by a few White doulas included the assumption that they had been hired by Black clients in the past because their clients felt it gave them greater authority in the hospital setting to have a White doula present in the room.

These points led doulas back to conversations surrounding the importance of the doula’s role, which generated **theme two**, with 100% of doulas reporting the importance of championing for *self-advocacy* while providing *advocacy* in the context of health disparities. Doulas reflected on prior instances of systemic racism they had witnessed their clients experience and commented on having to balance their role in these situations as an advocate while still needing to ensure access (particularly in the delivery room). They discussed the importance of educating about self-advocacy above simply providing *their own* advocacy. By teaching clients how to ask their own questions, represent their interests, and convey the needs and rights of their body and their child’s, doulas reported providing a skill set that could be passed on beyond the pregnancy. However, doulas did concede that self-advocacy is a skill that takes practice - and is “easier said than done.” Notably, the importance of advocacy was highlighted around labor and delivery. Doulas did mention White doulas needing to avoid a savior mentality with Black clients. Doulas reported the hospital setting as especially challenging (both for themselves and their Black clients), as many healthcare providers may not accept their presence, leading to complexities in being able to fulfill their role to their utmost potential in these settings.

Considering themes one and two, doula responses revealed multiple action steps for implementation. Proposed action steps (with quotes) in Table 3 are also linked to a coordinated SEM level for further exploration. First, doulas addressed (1) their personal responsibilities in being informed on racial disparities regarding maternal health disparities. Beyond “knowing the statistics,” discussions of having a responsibility to be aware of systemic racism were addressed. Some doulas discussed the importance of (2) having doula certification programs tie more systemic racism and implicit bias education into certification curricula so that this information is provided to more doula communities. Some doulas reported smaller organizations as being more inclusive of racial and ethnic diversity efforts. Therefore, some reported that larger organizations may not be as in tune with community needs or outreach, but that these organizations provide the pathway for helping more clients in-need. Many participants reflected diversity of doulas in their communities as an issue. Doulas reported a need for (3) greater public health funding to provide increased support for doula compensation and recruitment of diverse doulas to ensure clients have options. Finally, (4) some doulas recommended that the doula-provider communicative relationship be improved upon by partnering with local hospitals in substantive ways.

**Table 3.** Themes and Proposed Action Items for Utilizing Doulas to Mitigate Maternal Morbidity and Health Disparities.

| Theme | Relevant SEM Level(s) | Actionable Steps | Actionable Sub-Steps | Relevant Quote(s) |
| --- | --- | --- | --- | --- |
| (1) Doulas identified significant racial disparities for Black women when it comes to maternal morbidity and health disparities. | Public policy<br>Community<br>Institutional<br>Interpersonal<br>Individual | a) doulas have a personal and professional responsibility to be educated on obstetric implicit bias<br>b) national doula certification programs should more strategically integrate implicit bias into their curriculum in an effort to get this information dispersed into communities | 1) Ensure that curriculum components are developed based on prior work of scientists and birthing people from historically marginalized groups – and that any courses provided are taught by these individuals as well, to ensure an informed, patient-centered design in all phases of implementation<br>2) Proper message testing of all materials will be necessary (with member-checking and oversight by a health communication scholar) to ensure that the education being provided to doulas is culturally | *“We make sure we're creating a doula community that is culturally and linguistically, racially, ethnically, and religiously representative of birthing people. So making sure that we're doing that and, in the event that like Angela and I are both White we, you know, we can't change that. So making sure that Angela and I have done the work to remove our own biases to make sure that we're not bringing sort of that layered racism into the room, implicit bias and things like that, if we're working with clients that are anything but like White, English-speaking people. And so I think that's really important for doula to do that because we're that consistent support, we've been with you prenatally, were often with you during Labor, before you go to the hospital, we're there with you through triage, there through leave for nurses changing and doctor shifts changing, all that. We're that consistent person, so making sure that we've done all that we can to be the best consistent...yeah the best person there for you during that time and acknowledging all of that.” – Samantha*<br>*“...just insurances here in Florida, or probably nationwide, a very limited number of organizations are considered as qualified doulas, so if you haven't gone through one of the five major doula organizations, it doesn't matter, you're not going to contract with any insurances, because you didn't go through DONA, or you didn't go through CAPP, or you didn't go through ICEA, it's basically like 3-5 major ones. And the issue with that is that those organizations have their own systemic issues and problems as far as being inclusive, as far as being anti-racist, as far as being anti-trans, there's just a lot of issues in that respect. Also they are quite expensive. If you've gone through a smaller training organization that is maybe led by Black women or has a focus on trans or LGBT clients, then you may be getting the educational piece and the information you need to appropriately serve the* |
|  |  |  | relevant, sensitive, and socially responsible | *community you want to serve, but then you're lacking the access piece to where people can actually access you because you can't qualify to be a Medicaid provider, and so it severely limits the diversity of doulas that are available on Medicaid, which is like the biggest problem because you have that diversity with your Medicaid clients but you don't have the diversity with doulas, so that is a major barrier between doulas and the Medicaid system... ” – Katy* |
| (2) Doulas reported that they help to reduce maternal morbidity and health disparities for Black birthing people by promoting self-advocacy and providing advocacy when necessary. | • Interpersonal | a) greater public health funding is necessary to bolster the doula profession and to help diversify doulas for various racial and ethnic communities<br>b) local hospital partnerships should be established to begin working toward a “cycle of birthing communication ” aimed at breaking down any barriers that may exist between healthcare providers and doulas, | 1) Roundtable discussions between healthcare providers (nurses, OB-GYNs and doulas) to discuss their roles, with emphasis on shared goal of healthy birthing process<br>2) Having an expert in organizational communication present may be helpful to ensure that all conversations are constructive and positive | *“I would say the best scenario that I’ve seen tried to have a positive note here is that for quite a few years at [redacted]. There was a woman named [redacted], who was the head of DONA at one time, and she had a relationship with [redacted] and created like a [redacted] sort of. She was trying to create this bridge between doulas and like labor and delivery. And so, as a result, there would be like annual events where they would invite all these doulas and all these OBs and nurses and stuff like that, so it was like networking and then we’ve had these...Some of us we would get asked to these regular roundtable meetings where you know nurses would complain, would tell us like what they hated about doulas quite frankly, like, the shitty things that had happened, or like things that were upsetting to them, or like where we were stepping on toes and stuff like that, and then we could say “this is what I have an issue with,” like “the fact that I couldn't get into the OR,” or whatever, so there was an actual space for this. There was a few years there were there was a very open dialogue between doulas and labor delivery and it totally changed the tone of [redacted], it became my favorite place to go, like I could wear my advisory board badge and like all that kind of stuff you know. Unfortunately, it like unraveled at some point, there was some issue or whatever between [redacted] and [redacted] or one of the OBs I don't know I don't know what happened, I just know that it got disbanded but there was a while that having that open dialogue was a huge very positive thing you know and they did something similar at [redacted] but it wasn't quite nearly as vigorous or any of that kind of thing. But it was still created by like one of the directors of nursing and stuff like* |
|  |  | particularly in the hospital setting |  | *that, and so you know similar they would like invite like 16 of us to come and be in conversation, and they would tell us about things that they were trying to do on their end and you know request things of us and like stuff like that so...that was a big deal. That was a big deal. That is something that I can see really making a difference here in Florida.” - Jane* |
| (3) Doulas perceive their role in helping to reduce maternal morbidity and health disparities, by providing access to resources, information, and education. | • Community<br>• Interpersonal | a) doulas should provide tailored resources for their clients, based on demographic information, familial structure, and geographic location<br>b) partnering with hospitals with ensure that messaging is representative is recommended | 1) Resources may include:<br>o local providers<br>o birth plan worksheet<br>o lactation support groups<br>o screening tools for mental health<br>o local parenting classes<br>o list of pediatricians<br>o affirmation<br>o “when to call your doula”<br>2) Keep in mind when working with messaging that representation matters – but authentic representation is key; therefore, message testing is important to ensure patients of all backgrounds feel represented | *“...the types of materials or in the materials that you see provided, for example, in the hospital setting ....it’s just the issue of representation.... that’s really important. Yes, it shouldn’t always be, for example, a White mom. It’s really important that Black mothers can see Black mothers in some of these materials. Even just the standard medical textbooks and posters that you see in your provider’s office – it’s always going to be a White baby, even if it’s animated. Whenever you see a poster of a baby and a pelvis it’s going to be a White baby there. It is. I have never seen a baby of color on a poster in a doctor’s office, ever. Oh yeah, there’s a huge issue of representation.” - Chanel*<br>*“So that’s something that I had a conversation with my trainer about (my training for my certification). That’s something that they are working on because their standard packages that they have and those paper resources – they’re pretty basic as far as “mom and her partner” and the partner is always “dad.” You know, “the dad’s handbook” or “the dad’s car” – so, stuff like that, so yeah that’s something that I can tweak to cater to my client. I’m not going to hand them a packet that says “my partner can do this during labor; my partner can help me do this postpartum....” when they don’t have one.” – Chanel* |
|  |  |  | 3) every family structure looks different – provide different resources for each scenario |  |
| (4) Doulas engage in positive social surveillance – the act of consistent, open communication, to help keep a positive, watchful eye on their clients. | • Interpersonal<br>• Individual | a) check-in with clients on a regular basis<br>b) keep scope of practice in mind (referring clients back to healthcare providers for medical concerns) | 1) Most preferred forms of communication according to doulas:<br>o text<br>o email<br>o face-to-face<br>2) Check-in with clients after their appointments to see how things went and cultivate a positive doula-client relationship built on trust<br>3) Use positive social surveillance as a way to champion self-advocacy<br>4) But be another “set of eyes” for your client whenever possible | *“I think the relationship that you can have with your client and then maybe what you're doing in class and you look at them and see how swollen they are, you say “you know, I'm concerned, you need to check in with your midwife,” or chatting with them and seeing dark circles and them saying “I feel so tired,” ... ”you need check...” -- you don't want to say “need” – but, you know, “I highly recommend you check in with your midwife it's these signs are known to be when you're very anemic.” And so I think we can help with that and being in the hospital where maybe moms aren't being listened to you to help them to have that voice.” – Emma*<br>*“Staying with, especially hospital clients, and mother and baby for a couple hours immediately postpartum because the late onset postpartum hemorrhage is one of the biggest things that I have seen happen in a hospital setting post-delivery that, just due to the nature of having, to have more than, you know, with nurses being short staffed, especially now, and in and out of the room, they are not able to sit and watch your client as closely as you are. So staying with those clients for just a couple hours to watch them. Watch their body language, you know, help them go to the bathroom, help them do all of those things. It definitely helps. It's a second set of eyes that the hospital just can't provide due to the nature of the establishment, quite often. I still think that maternal morbidity falls within our LGBT and or BIPOC population, although I have seen it in White women as well, the levels of care are just not the same. I actually was discussing with my own personal therapist who has five kids (so she gets my level of crazy), and we were discussing birth trauma and PPD and PPA and she's telling me her story -as very educated Black woman how she was treated in New York. And I looked at her, and I said,* |
|  |  |  |  | *“all I can think of is if you'd been a White woman this wouldn't have happened. And it was a late onset postpartum hemorrhage and she needed a blood transfusion and she probably needed more than one unit and she only got one unit, it was just barely enough to get her by, and now we know that a milk is made from blood and she's having nursing complications because her blood stores were low, and you know it's this downward spiral Of unnecessary tragedy. She survived. Great. Yay. Congratulations. You're alive.” [sarcasm] - Kristen* |

When doulas were asked about how they perceived their role in helping to reduce maternal morbidity and maternal health disparities, two final themes strongly emerged. **Theme three** was classified as *education and resources*, where 92% of doulas described providing access to resources, information, and education. Often, these resources were discussed as relating to childbirth, with the doula helping the birthing person prepare for what to expect during labor. Provision of resources were deemed a necessity for the postpartum period. Additionally, many doulas discussed the importance of resources specific to a client’s individual needs and geographic area. A few doulas discussed the significant effort required to put together different versions of targeted resource packets for different client situations, but they did not underestimate the importance of these resources in helping their clients feel more informed.

Regarding *education and resources*, doulas reported that it was important that each doula was well-informed on their community and its providers and services (e.g., lactation consultation, mental health services, etc.) for a variety of client profiles (e.g., insured vs. Medicaid recipient). Therefore, a viable action step would be (5) the creation of customizable resources for various client situations, complete with culturally inclusive messaging. Partnering with existing local community organizations is vital. Some doulas commented that (6) a tailoring of resources in hospital settings may be necessary not only to ensure that clients from all populations, especially marginalized groups, are being represented, but to ensure they are being *meaningfully represented*, not featured in a tokenistic way.^29^

Finally, **theme four** revealed that 82% of doulas tied the importance of their role back to the act of consistent, open communication with their clients, classified as *positive social surveillance*. Such encounters involved doulas making themselves available for their clients via phone, text, or email and providing greater access than a healthcare provider might be able to. By checking in, listening, and having someone check-in on them after an appointment, doulas reported that the relationship can become stronger over time. Many doulas extended this theme by noting what or was not within their scope of practice, indicating that when a client asks a medical question, they refer to the healthcare provider. Therefore, they identified their role as one with the ability to effectively monitor for social situations and inequities that other members of the healthcare team may not be privy to due to their time constraints or lack of access to the client.

Action steps suggested for theme four include helping clients set reminders for appointment times, providing referrals for clients who may need greater levels of assistance based on various social determinants of health (particularly focusing on partnerships with racially and ethnically based community organizations when able), and in instances where implicit bias is perceived (via the act of positive social surveillance), reminding the client of their self-advocacy skills and acting as an advocate when necessary.

## Discussion

Our research found that participants overwhelmingly associated MM and SMM with Black or African American birthing people. Described racial disparities were attributed to systemic racism and implicit bias.^30, 31^ In addition to providing education, resources and emotional support, our participants discussed the importance of their role in assisting birthing persons and families by teaching self-advocacy and providing advocacy in the healthcare setting (SEM interpersonal/institutional level). Doulas assist birthing people in navigating their healthcare journeys, processing the healthcare field, overcoming obstacles, and providing valuable information. Where physicians can detect “high risk” situations in ways that doulas cannot, doulas may be able to detect “social high risk” in ways that physicians cannot, due to such acts of *positive social surveillance*. They appreciate their ability to assist birthing people with navigation of a complex healthcare delivery system that may hamper access to care (e.g., institutionalized formalities and protocols) (SEM institutional level).

Doula support during pregnancy, labor, and delivery has been associated with improved outcomes among women facing disproportionately high rates of maternal morbidity ^32^. Doula clients report higher rates of satisfaction with their birth experience and an increased belief in their own ability to influence birth outcomes through behavior change ^33^. Our previous work demonstrates Black women who are supported by doulas during pregnancy and postpartum report enhanced communication and information sharing as well as improved emotional support ^34^. These findings align with results of the current study which indicate that doulas feel their primary role is to support birthing persons during pregnancy, birth, and postpartum (SEM interpersonal level). Conversations focused on doulas having a responsibility to their communities by being aware of systemic racism and implicit bias, utilizing culturally representative educational efforts, and continuing to engage in positive surveillance by paying attention on behalf of their clients while not going outside their scope of practice.

Doula responses revealed multiple action steps for implementation (Table 3) including doula certification programs include more on systemic racism and implicit bias education and awareness (e.g., one suggestion for how to begin a foundation for this is Royce et al.’s Special Report on addressing implicit bias in obstetrics and gynecology education).^31^ Regarding need for improved doula-provider communicative relationship, we suggest working toward a “cycle of doula communication,” where there is a clearly defined goals and roles for all stakeholders from the beginning to avoid what may have been a historically adversarial relationship. Health communication experts could provide training programs, scripts, and suggestions for events ranging from monthly webinars/meet-and-greets and brown bag lunches between healthcare providers and doulas.

### Limitations

We acknowledge that a limitation of our study is our sample structure. While racially representative of the larger doula profession, it is important that our future work focus on purposive sampling that explore doulas’ perspectives from minoritized communities on their ability to provide support for clients, particularly in the context of medical racism. Considering our findings regarding a need for more Black doulas, it is necessary to understand how Black doulas feel they may influence MM/SMM or related disparities.

Additionally, an advantage that doulas may have over healthcare providers is their ability to engage with multiple facets of a patient’s behavior (e.g., diet, exercise, appointment compliance, testing, etc.), referenced earlier as positive surveillance. When discussing an issue as complex as SMM, doula support for each of these areas certainly play a role and were addressed in our interviews and focus groups but were beyond the scope of this study due to space limitations.

## Health Equity Implications

Overall, our results revealed that doulas feel they serve an important role in helping to address health inequities in the perinatal period. Many of their goals entail building their professional identity around ways to dismantle these preventable occurrences for socially disadvantaged clients. Notably, our study and its findings align with the Commonwealth Fund’s Advancing Health Equity Program. This study supports the foundation’s evidence-based approaches for equity-centered maternity care which included the importance of doulas.^20^

Our findings and proposed action steps also have important policy implications. President Biden’s 2023 budget includes $470 million toward investments (e.g., the Healthy Start Initiative) to expand maternal health initiatives aimed at improving access to care to address the crisis of Black maternal morbidity and mortality in the United States.^35^ Specifically, proposed calls to action from the Health Resources and Services Administration include the implementation of bias training for providers and the hiring, training, certifying, and compensation of community-based doulas in areas with higher rates of disproportionate health outcomes,^36^ which apply to the findings and recommendations of our study. As the state of Florida ranked 36 out of 50 in 2022 for the Commonwealth Fund’s scorecard for healthcare, we are at a crucial juncture where support services for families must be championed to help families survive, and ultimately, have the chance to thrive.

## Data Availability

All data produced in the present study are available upon reasonable request to the authors

## Acknowledgements

We thank Victoria Evans, MA (Clinical Research Coordinator at Pediatric Research Hub, University of Florida), for her editorial contributions.

## Authorship confirmation/contribution statement: Janelle Applequist

Methodology, Conceptualization, Software, Validation, Formal Analysis, Investigation, Data Curation, Writing – Original Draft, Writing – Review & Editing; **Roneé Wilson**: Conceptualization, Project administration, Funding acquisition, Supervision, Writing – Original Draft, Writing – Review & Editing; **Megan Perkins**: Validation, Investigation, Writing – Review & Editing; **Charlette Williams**: Conceptualization, Investigation, Writing – Original Draft; **Ria Joglekar**: Conceptualization, Investigation; **Richard Powis**: Conceptualization, Writing – Review & Editing; **Angela Daniel**: Conceptualization, Writing – Review & Editing; **Adetola F. Louis-Jacques**: Conceptualization, Project administration, Supervision, Writing – Original Draft, Writing – Review & Editing.

